# The sensitivity of respiratory tract specimens for the detection of SARS-CoV-2: A protocol for a living systematic review and meta-analysis

**DOI:** 10.1101/2020.07.02.20144543

**Authors:** Adam J. Moore, Maura I. Nakahata, Chaney C. Kalinich, Kate Nyhan, Daniel J. Bromberg, Xiaoting Shi, Albert I. Ko, Nathan D. Grubaugh, Arnau Casanovas-Massana, Anne L. Wyllie

**Affiliations:** Department of Epidemiology of Microbial Diseases, Yale School of Public Health, Yale University, New Haven, CT, USA; Department of Environmental Health Sciences, Yale School of Public Health, Yale University, New Haven, CT, USA; Harvey Cushing / John Hay Whitney Medical Library, Yale University, New Haven, CT, USA; Department of Social and Behavioral Sciences, Yale School of Public Health, Yale University, New Haven, CT, USA; Center for Interdisciplinary Research on AIDS, Yale University, New Haven, CT, USA

## Abstract

**Background:** Highly sensitive, non-invasive, and easily accessible diagnostics for Severe Acute Respiratory Syndrome Coronavirus 2 (SARS-CoV-2) are essential for the control of the Coronavirus Disease 2019 (COVID-19) pandemic. There is a clear need to establish a gold standard diagnostic for SARS-CoV-2 infection in humans using respiratory tract specimens.

**Methods:** Searches will be conducted in the bibliographic databases Medline, Embase, bioRxiv, medRxiv, F1000, ChemRxiv, PeerJ Preprints, Preprints.org, Beilstein Archive, and Research Square. Relevant government documents and grey literature will be sought on the FDA’s Emergency Use Authorizations website, the ECDC’s website, and the website of the Foundation for Innovative New Diagnostics. Finally, papers categorized as diagnosis papers by the EPPI Centre’s COVID-19 living systematic map will be added to our screening process; those papers are tagged with the diagnosis topic based on human review, rather than database searches, and thus this set of papers might include ones that have not been captured by our search strategy.

## Introduction

Highly sensitive, non-invasive, and easily accessible diagnostics for Severe Acute Respiratory Syndrome Coronavirus 2 (SARS-CoV-2) are essential for the control of the Coronavirus Disease 2019 (COVID-19) pandemic. Upon discovering that the pneumonia outbreak in the City of Wuhan in Hubei Province, China was caused by a novel betacoronavirus, medical professionals had to rely on diagnostic methods already in place for the detection for an emerging infectious disease with no established gold standard^1–3^. While various polymerase chain reaction (PCR) methods are currently being used to test for the presence of SARS-CoV-2 viral RNA^4,5^, there is still no universally agreed upon gold standard for which diagnostic specimen should be collected, nor how it should be collected.

Nasopharyngeal (NP) and oropharyngeal (OP) swabs are considered to be the gold standard for specimen collection for the majority of upper respiratory tract infections^6,7^. The United States Center for Disease Control and Prevention had originally recommended NP swabs be used to collect a specimen for SARS-CoV-2 testing; however that recommendation has now been updated to include OP swabs, nasal mid-turbinate (NMT) swabs, anterior nasal (AN) swabs, NP wash/aspirate, and nasal wash/aspirate^8^. Incidentally, NP and nasal swabs have demonstrated insufficient sensitivity for detecting SARS-CoV-2 in various studies, particularly amongst asymptomatic patients and those with low viral loads^9–13^. Saliva is perhaps the only clinical upper respiratory sample type still not formally recommended by the US CDC, despite showing promise for early viral RNA detection while circumventing the supply chain bottlenecks all swabs are subject to and having received Emergency Use Authorization (EU) by the US Food and Drug Administration (FDA) in at least once testing facility at Rutgers University^8,12,14^. As such, clinicians are left with too many choices and not enough guidance on which specimens should be collected and how collection should be carried out.

There is a clear need to establish a gold standard diagnostic for SARS-CoV-2 infection in humans using respiratory tract specimens. We are aware of other, currently on-going and completed reviews of a similar nature, but none are comparing all of the specimen collection methods we are analyzing (NP, NMT, OP, AN swabs, NP and nasal washes, sputum, and saliva) nor do they implement the extensive search we show in this project^15–19^. We are implementing a comprehensive and rigorous search and analysis strategy, with the goal of providing clear guidance to clinicians on which specimen type and collection method is preferred for accurate detection of SARS-CoV-2. The initial results will be published in a manuscript, following the PRISMA reporting guidelines^20^. Monthly updates will occur for the duration of 2020 and be made available on the web. After the final web update, the published manuscript will be updated to reflect the data collected and analyzed since publication.

### Primary Questions

1. Which respiratory tract sample collection method is the most sensitive and specific for the detection of SARS-CoV-2 in humans?
2. How do the sensitivity and specificity of detection for each specimen type compare to one another?

### Domain of study

Studies that report the sensitivity of SARS-CoV-2 upper respiratory specimen diagnostics and specimen collection methods.

### Searches

The search strategy will be designed by a medical librarian (KN) and peer reviewed by an independent medical librarian. The search has two elements: search terms related to sampling methods and search terms related to COVID-19. These two concepts will be operationalized with controlled vocabulary and keywords, drawing on existing search strategies, including the Ovid Expert Search for coronavirus. Potential expansions will be investigated: adding OR statements for diagnostic test performance and adding OR statements for screening. The searches will be updated shortly before the manuscript is submitted and again once per month for the duration of 2020.

Searches will be conducted in the bibliographic databases Medline and Embase (both on the Ovid platform). For the final update, a search may be conducted in PubMed, to take advantage of PubMed’s earlier receipt of article metadata from publishers.

Relevant preprints will be sought. Those with NIH funding may be retrieved through our MEDLINE search due to PubMed’s recent inclusion of NIH-funded preprints, but more will be sought through EuropePMC, which indexes preprints from bioRxiv, medRxiv, F1000, ChemRxiv, PeerJ Preprints, Preprints.org, Beilstein Archive, and Research Square.

Relevant government documents and grey literature will be sought on the FDA’s Emergency Use Authorizations website, the ECDC’s website, and the website of the Foundation for Innovative New Diagnostics.

In addition to database searching, the references in included papers and more recent publications citing included papers will be reviewed for additional relevant documents (backwards and forward citation chaining). Similarly, the references in similar reviews will be reviewed.

Finally, the papers categorized as diagnosis papers by the EPPI Centre’s COVID-19 living systematic map will be added to our screening process; those papers are tagged with the diagnosis topic based on human review, rather than database searches, and thus this set of papers might include ones that have not been captured by our search strategy^21^.

Records that meet the following criteria will be move into the full-text screening:

1. Published in either English, Spanish, French, Italian, Portuguese, or Chinese.
2. Published in a peer-reviewed journal or available on the web from a preprint server, governmental sources, or non-profit organization.
3. Published in 2020.
4. Focused on human subjects who are, or are suspected of being, infected with SARS-CoV-2.
5. Provide a direct indication or otherwise imply PCR-based diagnostic testing was performed on the patients

Records that meet the following criteria will not be included in the full-text screening:

1. Published in a language other than English, Spanish, French, Italian, Portuguese, or Chinese.
2. A commentary/opinion piece.
3. Focused on non-human patients.
4. Explicitly states they only use non-PCR based diagnostics.
5. Only uses non-respiratory samples for diagnostics.

In order to be included in this systematic review, all records must meet the following criteria:

1. Published in either English, Spanish, French, Italian, Portuguese, or Chinese.
2. Published in a peer-reviewed journal or available on the web from a preprint server, governmental sources, or non-profit organization.
3. Published in 2020.
4. Focused on human patients who are, or are suspected of being, infected with SARS-CoV-2.
5. Research methodologies pertaining to specimens collected, the method of collection, tools used for collection, laboratory tests run on specimen(s), analysis of test results, and reporting are provided and clearly described in order to undergo quality assessment.
6. Include a comparison of PCR-based diagnostics performed on at least two different respiratory specimens.

Records will be excluded from the review if they meet the following criteria:

1. Studies that do not include paired comparisons between two PCR-based diagnostics on respiratory tract specimens.
2. Records that do not provide detail on the specimens tested and/or how they were collected.

### Draft search strategy (showing potential expansions)

#### Ovid MEDLINE(R) ALL <1946 to June 25, 2020>

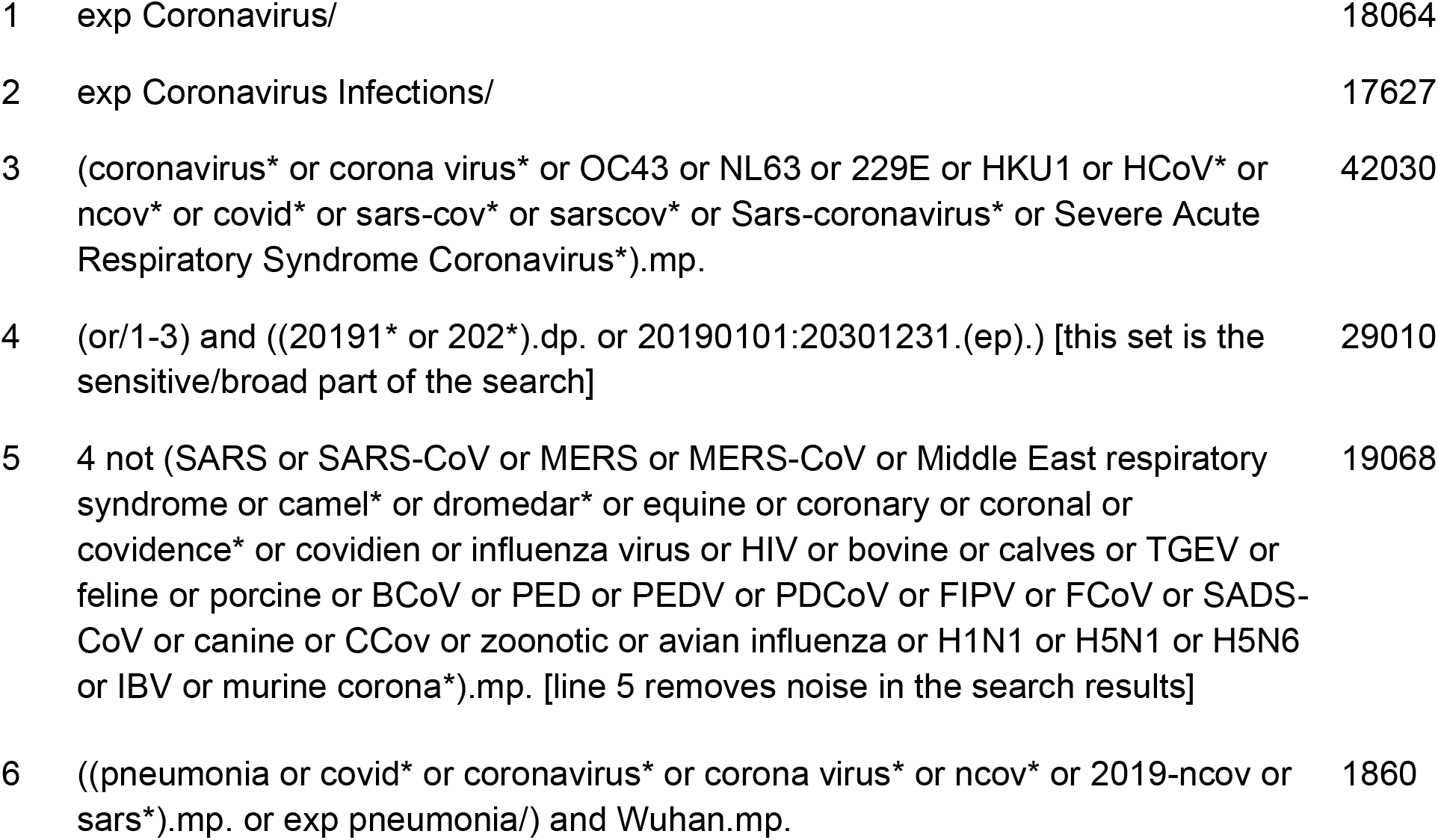

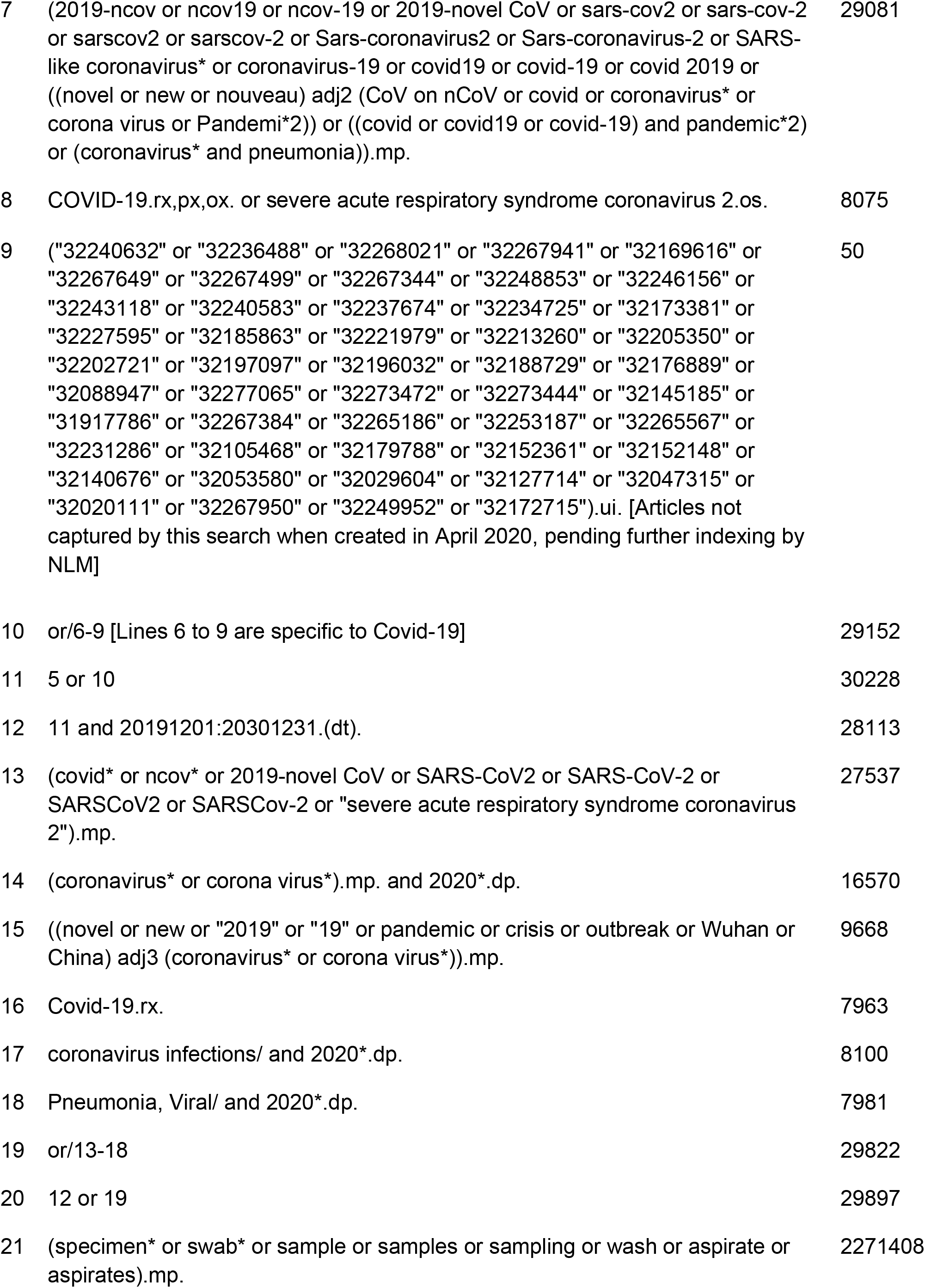

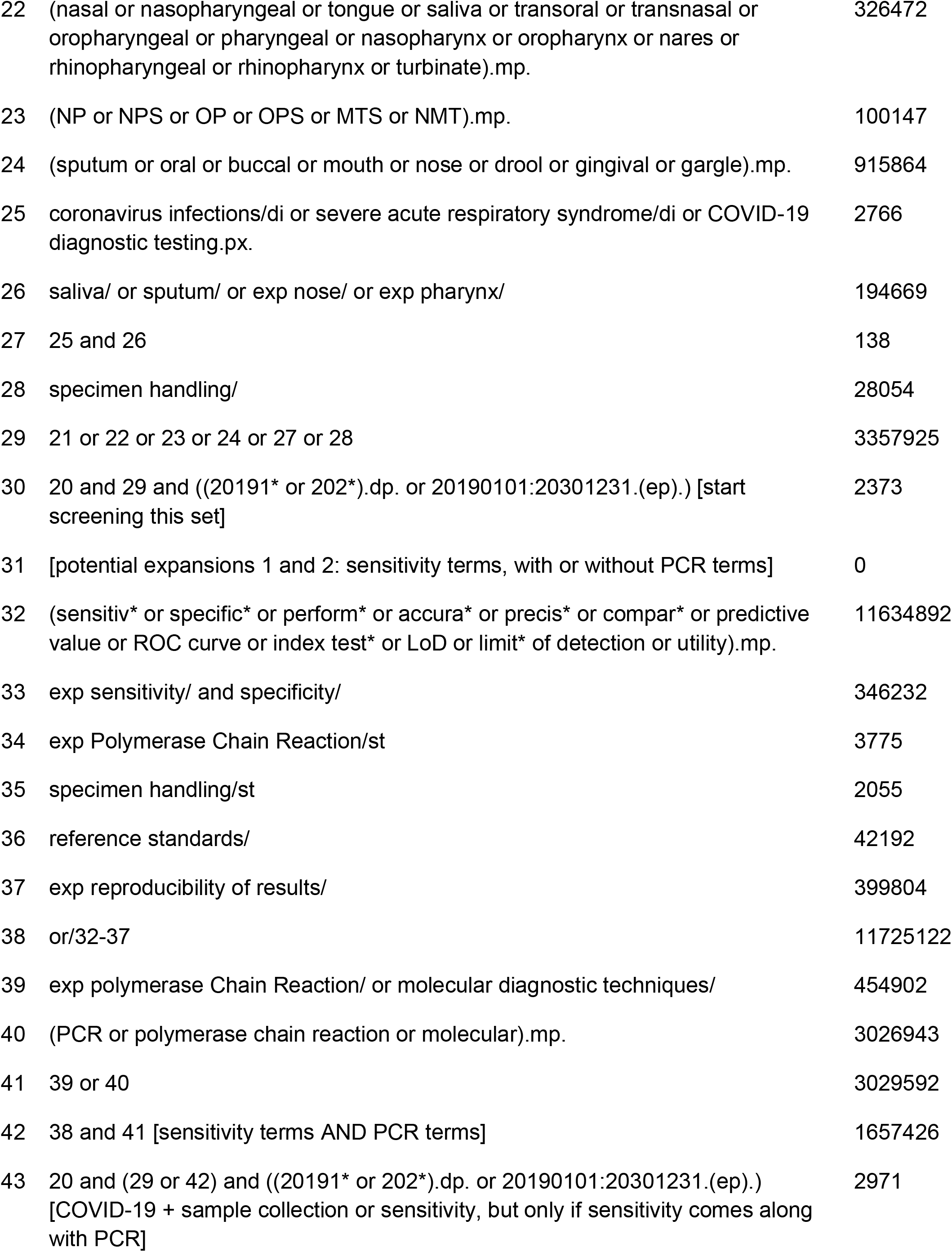

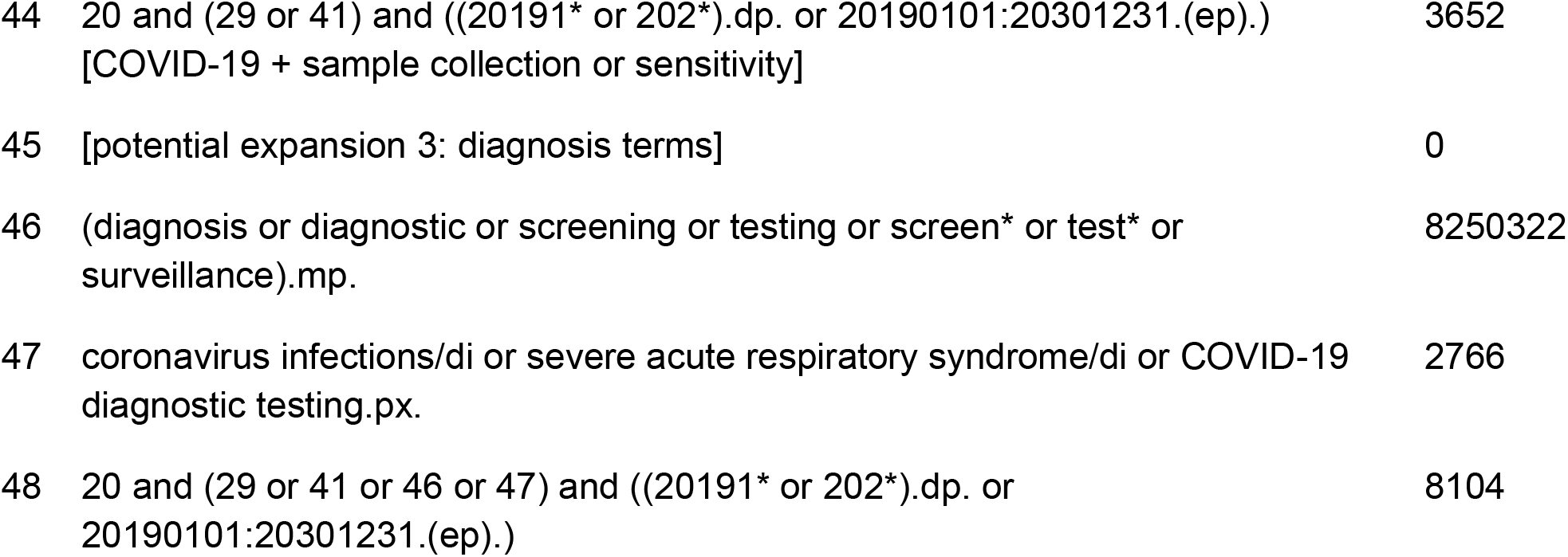

### Types of study to be included

This review will consider records that describe methods of respiratory tract specimen collection for the purpose of SARS-CoV-2 viral RNA identification by PCR. Peer-reviewed and preprint case studies, case-control studies, cohort studies, observational studies, assay validation studies, and US and European government documents in English, Spanish, Italian, French, Portuguese, and Chinese will be included.

### Participants/population

Individuals who have been tested for SARS-CoV-2 infection.

### Comparator(s)/controls

Respiratory tract specimens collected for SARS-CoV-2 viral RNA detection.

### Main outcome(s)

Sensitivity and reproducibility of specimen collection methods in the general population with SARS-CoV-2 infection.

#### Measure of effect

Sensitivity, specificity

### Data extraction (selection and coding)

We will be following the Joanna Briggs Institute (JBI) Reviewer’s Manual^22^. Primary data selection and extraction will be performed in Covidence. In both the title-abstract and full-text screening stages, each record will be screened by two co-authors independently (except for records in Spanish, Italian, French, Portuguese, which will be screened by AC-M and records in Chinese by XS).

After search results are uploaded to Covidence, co-authors AJM, MIN, CCK, and ALW will screen the title and abstract of all retrieved English articles and documents to identify those that might match the aforementioned inclusion criteria. Records that clearly do not meet the criteria will be eliminated at this stage. Those that remain will proceed to full-text review. Any conflicts in the title/abstract screening will be resolved by consensus or the involvement of a fifth co-author if necessary.

Records that appear to potentially meet the inclusion criteria at the title/abstract screening phase will then be subject to full-text review by co-authors AJM, MIN, and CCK to determine if they meet the inclusion criteria for this review. At the full-text screen stage, records that do not meet the inclusion criteria will be eliminated and the reason for exclusion will be logged. Conflicts on records that should be ex/included will be resolved with consensus between AJM, MIN, and CCK. We will perform a dual extraction of details on the study type, specimen type, method of specimen collection, true positive N, false positive N, false negative N, true negative N, total number of patients included, patient age and sex, primer/probe sets used for the PCR, and positive/negative cut off values. We will also extract the first author’s last name, the year of publication, country of publication, and title of the record. In cases where a preprint and a published article describe the same research, data extraction and risk of bias assessment will be performed on the published article. We will be following the reporting guidelines outlined by PRISMA(source). After the initial manuscript is published, the subsequent monthly searches and screenings will only be done on English language records.

### Risk of bias (quality) assessment

Each record that has been determined to meet the inclusion criteria upon full-text screening will be critically evaluated for risk of bias and reporting quality by at least two co-authors (AJM, MIN, CCK), following the risk of bias (critical appraisal) tools provided by JBI and the Standard for Reporting of Diagnostic Accuracy (STARD) checklist^22,23^.

### Strategy for data synthesis

Hierarchical modeling is the gold standard of meta-analysis of diagnostic test accuracy studies^24^. χ^2^ tests will be performed to assess the heterogeneity of sensitivities and specificities of tests in the sample. The sensitivity and specificity of all studies will then be meta-analyzed using the hierarchical summary receiver operating characteristic (HSROC) model. Analysis will be conducted with the *mada* package in R Studio^25,26^. Pooled sensitivity and specificity estimates, as well as 95% confidence intervals, will be calculated.

### Analysis of subgroups or subsets

If heterogeneity is found or suspected, a subgroup analysis will be performed as needed. Potential sources of heterogeneity include anatomical subsites such as mid-turbinate and anterior nasal areas, and demographic factors. If statistical pooling is not possible for these subgroups, the results will be presented in a narrative format.

### Anticipated or actual start date

1 July 2020

### Anticipated completion date

25 August 2020

## Data Availability

This is a protocol for a systematic review and there is no original data available at the time of this submission.

## Funding sources/sponsors

DJB has received funding from the National Institutes of Health (T32 MH20031). X.S. received a scholarship from China Scholarship Council. AC-M and AIK have received funding through research agreements with Regeneron. ALW has received research funding through grants from Pfizer and the National Basketball Association to Yale and has received consulting fees for participation in advisory boards for Pfizer.

## Conflicts of interest

The authors have no conflicts of interest to declare.

## Language

English

## Country

United States of America

## Stage of review

Review Ongoing

## Date of registration in PROSPERO

1 July 2020

## Details of any existing review of the same topic by the same authors

None

| Stage | Started | Completed |
| --- | --- | --- |
| Preliminary searches | Yes | No |
| Piloting of the study selection process | Yes | No |
| Formal screening of search results against eligibility criteria | No | No |
| Data extraction | No | No |
| Risk of bias (quality) assessment | No | No |
| Data analysis | No | No |

*The record owner confirms that the information they have supplied for this submission is accurate and complete and they understand that deliberate provision of inaccurate information or omission of data may be construed as scientific misconduct*.

*The record owner confirms that they will update the status of the review when it is completed and will add publication details in due course*.

## Notes

### Competing Interest Statement

The authors have declared no competing interest.

### Author Declarations

This is a systematic review that did not require IRB approval.

## References

1. Zhou P, Yang X-L, Wang X-G, et al. A pneumonia outbreak associated with a new coronavirus of probable bat origin. Nature. 2020;579(7798):270–273. doi:10.1038/s41586-020-2012-7

2. Wu F, Zhao S, Yu B, et al. A new coronavirus associated with human respiratory disease in China. Nature. 2020;579(7798):265–269. doi:10.1038/s41586-020-2008-3

3. Gorbalenya AE, Baker SC, Baric RS, et al. The species Severe acute respiratory syndrome-related coronavirus?: classifying 2019-nCoV and naming it SARS-CoV-2. Nature Microbiology. 2020;5(4):536–544. doi:10.1038/s41564-020-0695-z

4. Vogels CBF, Brito AF, Wyllie AL, et al. Analytical Sensitivity and Efficiency Comparisons of SARS-COV-2 QRT-PCR Assays. medRxiv, 2020. doi:10.1101/2020.03.30.20048108

5. Corman VM, Landt O, Kaiser M, et al. Detection of 2019 novel coronavirus (2019-nCoV) by real-time RT-PCR. Euro Surveill. 2020;25(3). doi:10.2807/1560-7917.ES.2020.25.3.2000045

6. Kim C, Ahmed JA, Eidex RB, et al. Comparison of Nasopharyngeal and Oropharyngeal Swabs for the Diagnosis of Eight Respiratory Viruses by Real-Time Reverse Transcription-PCR Assays. PLoS One. 2011;6(6). doi:10.1371/journal.pone.0021610

7. Kompanikova J, Zumdick A, Neuschlova M, Sadlonova V, Novakova E. Microbiologic Methods in the Diagnostics of Upper Respiratory Tract Pathogens. Clinical Research and Practice. 2017;1020:25–31. doi:10.1007/5584_2017_10

8. CDC. Information for Laboratories about Coronavirus (COVID-19). Centers for Disease Control and Prevention. Published February 11, 2020. https://www.cdc.gov/coronavirus/2019-ncov/lab/guidelines-clinical-specimens.html

9. Zou L, Ruan F, Huang M, et al. SARS-CoV-2 Viral Load in Upper Respiratory Specimens of Infected Patients. New England Journal of Medicine. 2020;382(12):1177–1179. doi:10.1056/NEJMc2001737

10. Wölfel R, Corman VM, Guggemos W, et al. Virological assessment of hospitalized patients with COVID-2019. Nature. 2020;581(7809):465–469. doi:10.1038/s41586-020-2196-x

11. Callahan C, Lee R, Lee G, Zulauf KE, Kirby JE, Arnaout R. Nasal-Swab Testing Misses Patients with Low SARS-CoV-2 Viral Loads. medRxiv. 2020:2020.06.12.20128736. doi:10.1101/2020.06.12.20128736

12. Wyllie AL, Fournier J, Casanovas-Massana A, et al. Saliva is more sensitive for SARS-CoV-2 detection in COVID-19 patients than nasopharyngeal swabs. medRxiv. 2020. doi:10.1101/2020.04.16.20067835

13. Tang Y-W, Schmitz JE, Persing DH, Stratton CW. Laboratory Diagnosis of COVID-19: Current Issues and Challenges. McAdam AJ, ed. Journal of Clinical Microbiology. 2020;58(6). doi:10.1128/JCM.00512-20

14. New Rutgers Saliva Test for Coronavirus Gets FDA Approval. Accessed June 22, 2020. https://www.rutgers.edu/news/new-rutgers-saliva-test-coronavirus-gets-fda-approval

15. Carver C, Jones, N. Comparative accuracy of oropharyngeal and nasopharyngeal swabs for diagnosis of COVID-19. Centre for Evidence-Based Medicine, Nuffield Department of Primary Care Health Sciences, University of Oxford. 2020.

16. Deeks J, Dinnes J, Davenport C, et al. Rapid point-of-care tests for diagnosis of SARS-CoV-2 infection. Cochrane Rapid Review. 2020.

17. Bwire GM, Njiro BJ. Detection profile of SARS-CoV-2 in different types of clinical specimens using polymerase chain reaction: a systematic review and meta-analysis. PROSPERO. 2020.

18. Paranhos LR, Siqueira W, Sabino-Silva R, et al. Salivary SARS-CoV-2 viral RNA for the diagnosis of COVID-19 patients: a systematic review and meta-analysis of the validity of the test. PROSPERO. 2020.

19. Azman M, Gendeh HS, Lum SG, Baki MM. Upper respiratory tract sampling in COVID-19. 2020:13.

20. Liberati A, Altman DG, Tetzlaff J, et al. The PRISMA Statement for Reporting Systematic Reviews and Meta-Analyses of Studies That Evaluate Health Care Interventions: Explanation and Elaboration. PLOS Medicine. 2009;6(7):e1000100. doi:10.1371/journal.pmed.1000100

21. COVID-19: Living systematic map of the evidence. Accessed July 1, 2020. <http://eppi.ioe.ac.uk/cms/Projects/DepartmentofHealthandSocialCare/Publishedreviews/COVID-19Livingsystematicmapoftheevidence/tabid/3765/Default.aspx>

22. Campbell J, Klugar M, Ding S, et al. Chapter 9: Diagnostic test accuracy systematic reviews. In: JBI Reviewer’s Manual. 4th ed. JBI; 2019. doi:10.46658/JBIRM-17-07

23. Bossuyt PM, Reitsma JB, Bruns DE, et al. STARD 2015: an updated list of essential items for reporting diagnostic accuracy studies. BMJ. 2015;351. doi:10.1136/bmj.h5527

24. Lee J, Kim KW, Choi SH, Huh J, Park SH. Systematic Review and Meta-Analysis of Studies Evaluating Diagnostic Test Accuracy: A Practical Review for Clinical Researchers-Part II. Statistical Methods of MetaAnalysis. Korean J Radiol. 2015;16(6):1188–1196. doi:10.3348/kjr.2015.16.6.1188

25. RStudio Team (2020). RStudio: Integrated Development for R. RStudio, PBC

26. Doebler P, Holling H. Meta-Analysis of Diagnostic Accuracy with Mada. 2015.

